# Triage-based care of people with Back Pain: STarT Back or Start diagnosing? An observational study

**DOI:** 10.1101/19005041

**Authors:** Tim Germon, Alex Jack, Jeremy Hobart

## Abstract

**Objectives:** Back pain is a massive public health problem. The STarT Back Screening Tool (SBST) was developed for use in primary care to triage people with lumbar pain, classifying them as low, medium or high “risk” of prolonged symptoms. This classification guides non-surgical interventions including manual treatments, exercise and cognitive behavioural therapy. Claims suggest SBST brings generic health and cost benefits. National guidance recommends STarT Back is used at the first primary care consultation but can be used at any stage. For SBST to be an effective triage tool it should distinguish structural from non-structural pain. We tested this requirement in consecutive people referred to a single triage practitioner, hypothesising it was not possible conceptually.

**Design:** An observational study of the relationship between routine, prospectively collected triage data and diagnosis.

**Setting:** A secondary care spinal triage service based in a teaching hospital.

**Participants:** We studied consecutive referrals with lumbar pain triaged by a single extended scope practitioner (ESP) over 22 months (Nov 2015-Sept 2017).

**Main Outcome Measures:** SBST and pain visual analogue scores (VAS: 0-10) were collected at the initial consultation. We compared data for people with and without surgically remedial lesions.

**Results:** 1041 people were seen (61% female, mean age 53), n=234 (28%) had surgically amenable explanations for pain. People with surgical lesions were older (58 v 51yrs), more likely male (48 v 35%) and had higher VAS scores (6.8 v 6.1). Surgery and non-surgery subgroups had similar SBST total and domain score distribution profiles. The surgery subgroup had less low risk (9%v21%) and more high risk (37% v 30%) classified people.

**Conclusion:** SBST scores did not differentiate surgical from non-surgical pathologies. It seems unlikely that symptom questionnaires can estimate prognosis accurately unless everyone has the same diagnosis, not just the same symptom. Diagnosis, rather than questionnaire scores, should guide treatment and inform prognosis.

**Summary Box**

What is already known on this topic?
The symptom of low back pain is a common cause of disability worldwide. The majority if people with low back pain probably do not have a structural problem in their lumbar spine to explain the extent of their disability. National guidelines throughout the world attempt to facilitate the identification and treatment of people with, “non-specific low back pain”, and prescribe treatment for people given this label.

What this study adds?
It seems unlikely that symptom questionnaires can estimate prognosis accurately unless everyone has the same diagnosis, not just the same symptom.
The practice of recommending treatment and prognosticating in the absence of a diagnosis needs further scrutiny.

## Introduction

Back pain is a huge and increasing public health problem. Estimates, published nearly two decades ago, suggested low back pain affects 24-36% of adults, persisted for ≥12 months 60-80% of the time, and that 85% of cases were “non-specific” (NSLBP).^1,2,3^ That is, more than 8/10 people with low back pain seeing primary care physicians were assumed to have no known specific underlying pathology or disease explaining their symptoms. Challenged health services responded by seeking management guidelines and methods to triage care.

The STarT Back Screening Tool (SBST; Fig 1) sought to address this need.^4^ SBST developers state: “there is a need for an adequately validated screening tool that allocates and prioritizes treatment for the entire spectrum of patients with NSLBP presenting to primary care, on the basis of treatment modifiable indicators, that is brief and quick to score”. Specifically, SBST aims to classify people with NSLBP as high, medium or low risk of prolonged symptoms and guide interventions (table 1). This approach is claimed to bring generic health and cost benefits.^5^

**Table 1.**
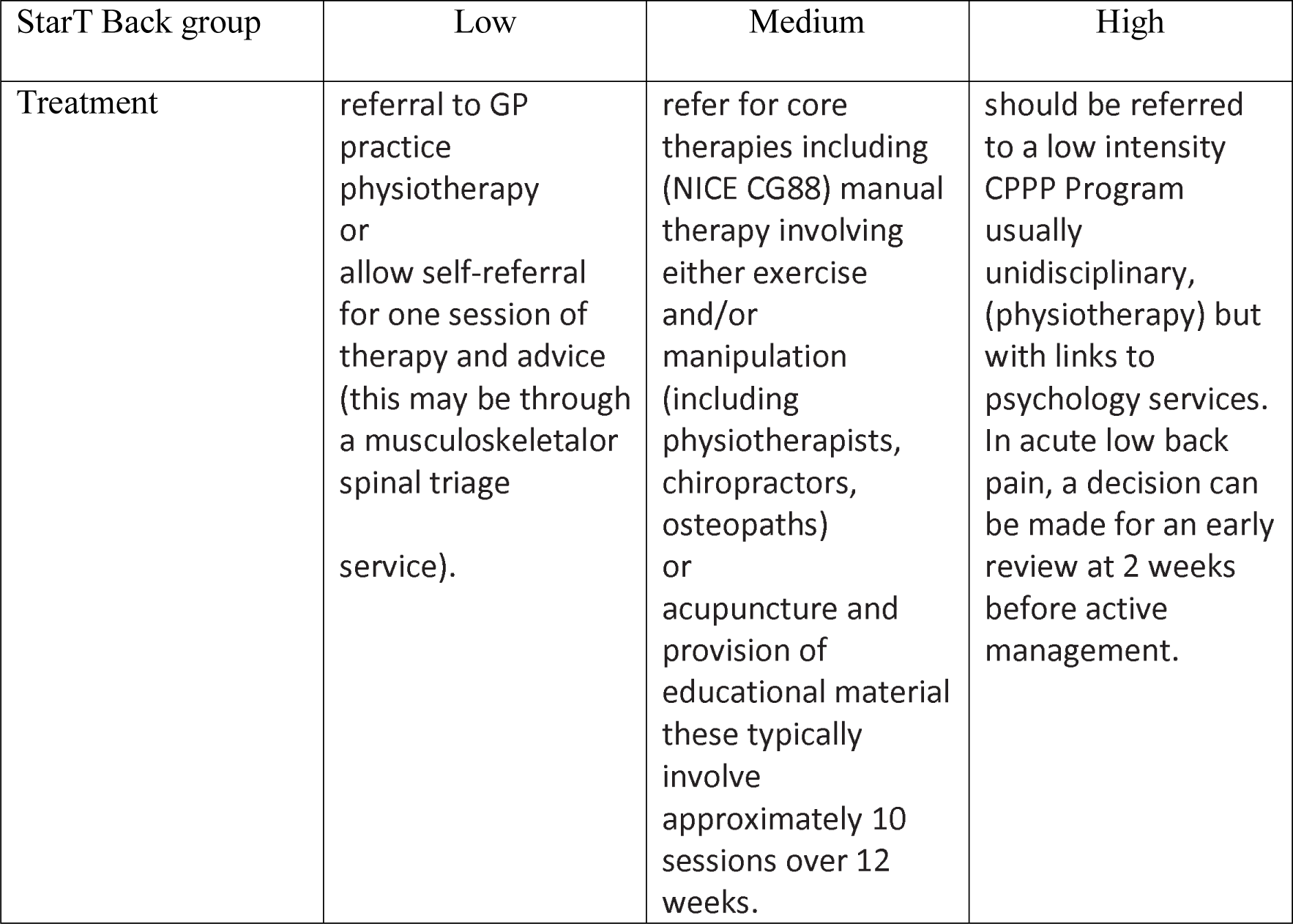
Treatment recommendations for risk groups according to Uk commissioning guidance.

**Figure 1.**
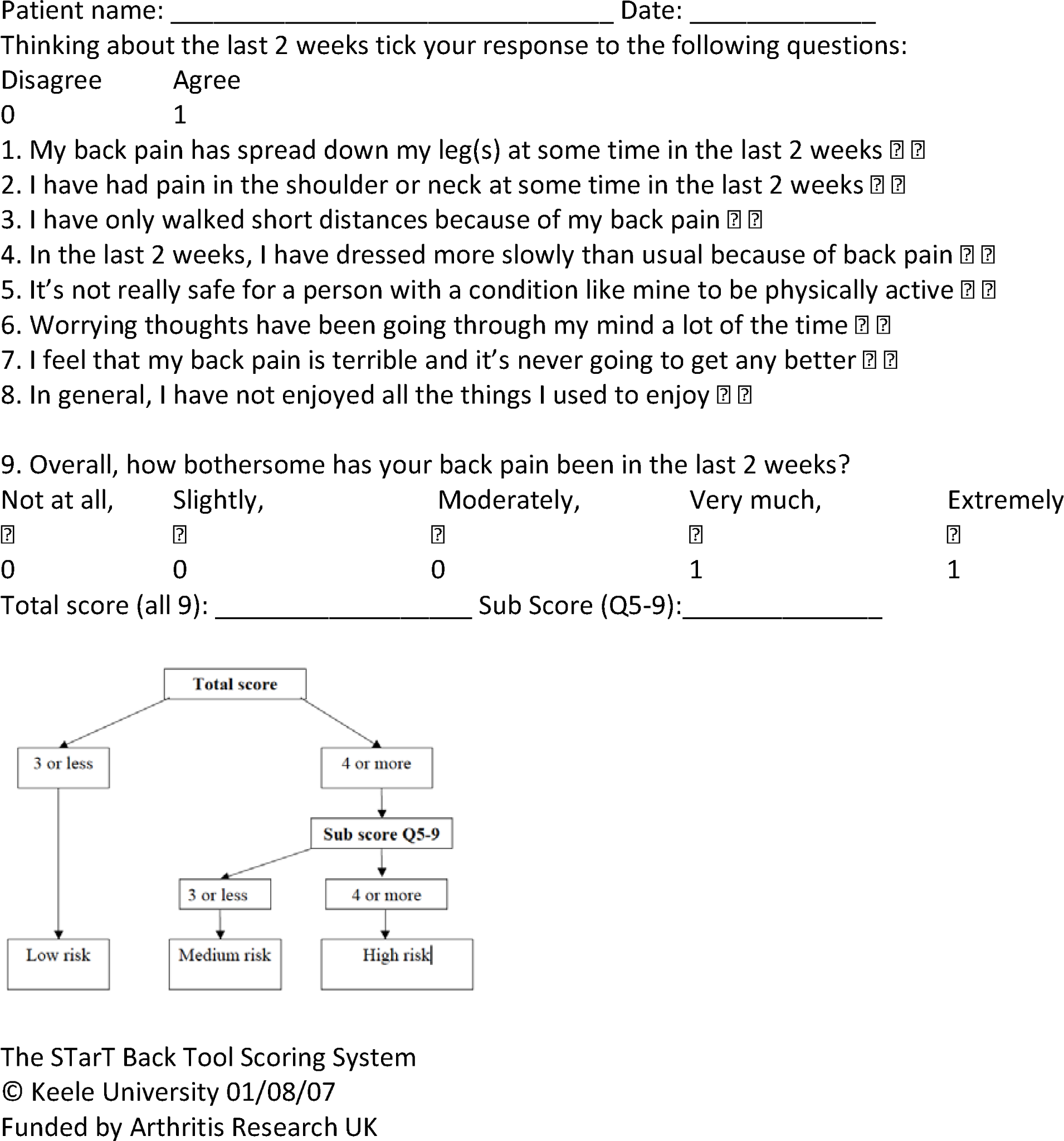
The Keele STarT Back Screening Tool.

Whilst SBST’s original focus on NSLBP appears clear, subsequent studies imply wider application to people with both specific and non-specific back pain.^4,5^ This suggestion is propagated in national guidance. Both the National Back & Radicular Pain Pathway and the British Orthopaedic Association (BOA) and The Royal College of Surgeons of England (RCSE) guidance to commissioners recommend SBST is administered to every person with back pain at their first primary care, as well as subsequent secondary care consultations (Fig 2).^6,7^

**Fig 2.**
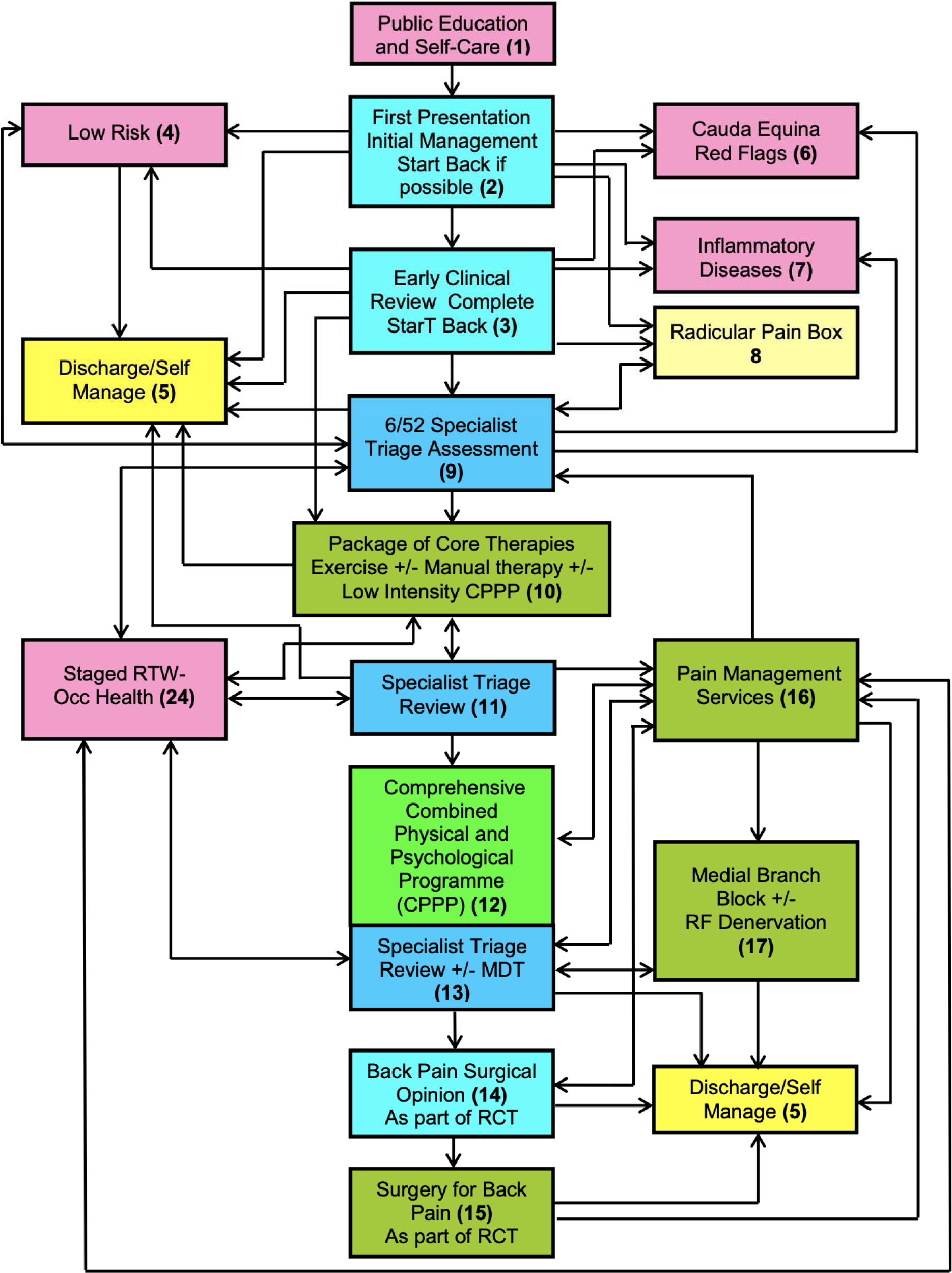
NHS England National Pathfinder Projects; **National Pathway of Care for Low Back and Radicular Pain, summary Page 14**.

This approach concerns us. First, it seems surprising to us that almost everyone with a common symptom has no specific cause. Second, a conclusion that symptoms have no explanatory cause is a diagnosis of exclusion requiring potential causes being excluded actively. This can’t necessarily be undertaken easily in primary care. Third, we have concerns about using symptom severity questionnaires to determine treatments in circumstances where the primary symptom, here pain, is the final common pathway of multiple diagnoses. Fourth, NSLBP is a label or description but not a diagnosis. We think this distinction is essential. Conceptually, people with one diagnosis could receive the same treatment, within reason. However, people with one label cannot be assumed to have one diagnosis. The only things common to people with “NSLBP” are pain and no clear explanation. Finally, we hypothesize that widespread application of SBST in primary care will triage people with treatable causes away from beneficial interventions. As such, diagnoses and beneficial treatments will be delayed or never realised.

For SBST to be used to identify risk, and guide interventions in primary care, a requirement is the ability to differentiate between surgical and non-surgical causes. We tested this requirement, hypothesising it was not possible conceptually for the reasons identified above.

## Methods

### Keele STarT Back Screening Tool (SBST; Figure 1)

The SBST is a self-report questionnaire for people with back pain. There are 9 questions (items). Eight items have two response categories: disagree and agree, scored 0 and 1. The ninth item has five response categories: not at all, slightly, moderately, very much, extremely; scored 0, 0, 0, 1, 1.

The SBST generates two scores. SBST total scores are generated by summing scores for all 9 items and range from 0 to 9. Higher total scores imply greater impact of back pain. SBST psychosocial subscale scores are generated by summing scores of questions 5-9 and range from 0-5. Higher subscale scores imply greater psychosocial impact.

The SBST score algorithm (Fig 1) defines people’s risk of persisting symptoms. First, SBST total scores define people’s risk as low (SBST total score </=3 [of max 9]), or not low (>/=4 [of max 9]). Next, the SBST psychosocial subscale scores define the not-low-risk group as medium risk (</=3 [of max 5]) or high risk (>/=4 [of max 5]).

### Sample

We studied all people referred for opinions regarding lumbar pain triaged by a single (AJ) extended scope practitioner (ESP) between November 2015 and September 2017. At the initial consultation the ESP made a clinical assessment and recorded SBST and visual analogue scale (VAS) pain scores (0 [no pain] to 10 [worst pain imaginable]). In accordance with our usual management, people thought to have clinical presentations suggesting structural abnormalities, principally pain thought to be indicative of nerve root compression, were offered MRI scanning. This is at odds with national guidance which only recommends MRI scanning prior to application of the STarT Back stratification in people with, “severe radicular pain not responding to treatment after 6-8 weeks”. Cases where scans are reported as suggesting lesions responsible for clinical presentations were discussed with consultant surgeons from the combined neuro-orthopaedic spinal service. When the surgeon considered the MRI abnormality explained the clinical presentation people were offered surgical assessments. In other words, people in our series with whom surgery was discussed would not have been offered that opportunity had national guidance been adhered to strictly.

### Analyses

We computed: baseline demographics, proportions of people who had MRI scanning and surgery, baseline SBST and VAS score distributions and summary statistics for people with and without surgically remedial lesions.

### Patient & Public Involvement

This is a retrospective analysis of routinely collected patient and clinician reported information as recommended by guidelines. In this context public and patient involvement in the design would not have been possible.

## Results

A total of n=1041 people were assessed by the ESP. Of these, n=489 (46.9%) had lumbar MRI scans of which n=137 (28%) had MRI-determined surgical lesions.

Table 2 shows the total sample mean age was 53 years (SD=16.4; range 16-91), 61.2% (n=638) were female. SBST data were available for 82.4% of people assessed. Missing SBST data were higher in the non-surgical subgroup.

**Table 2:**
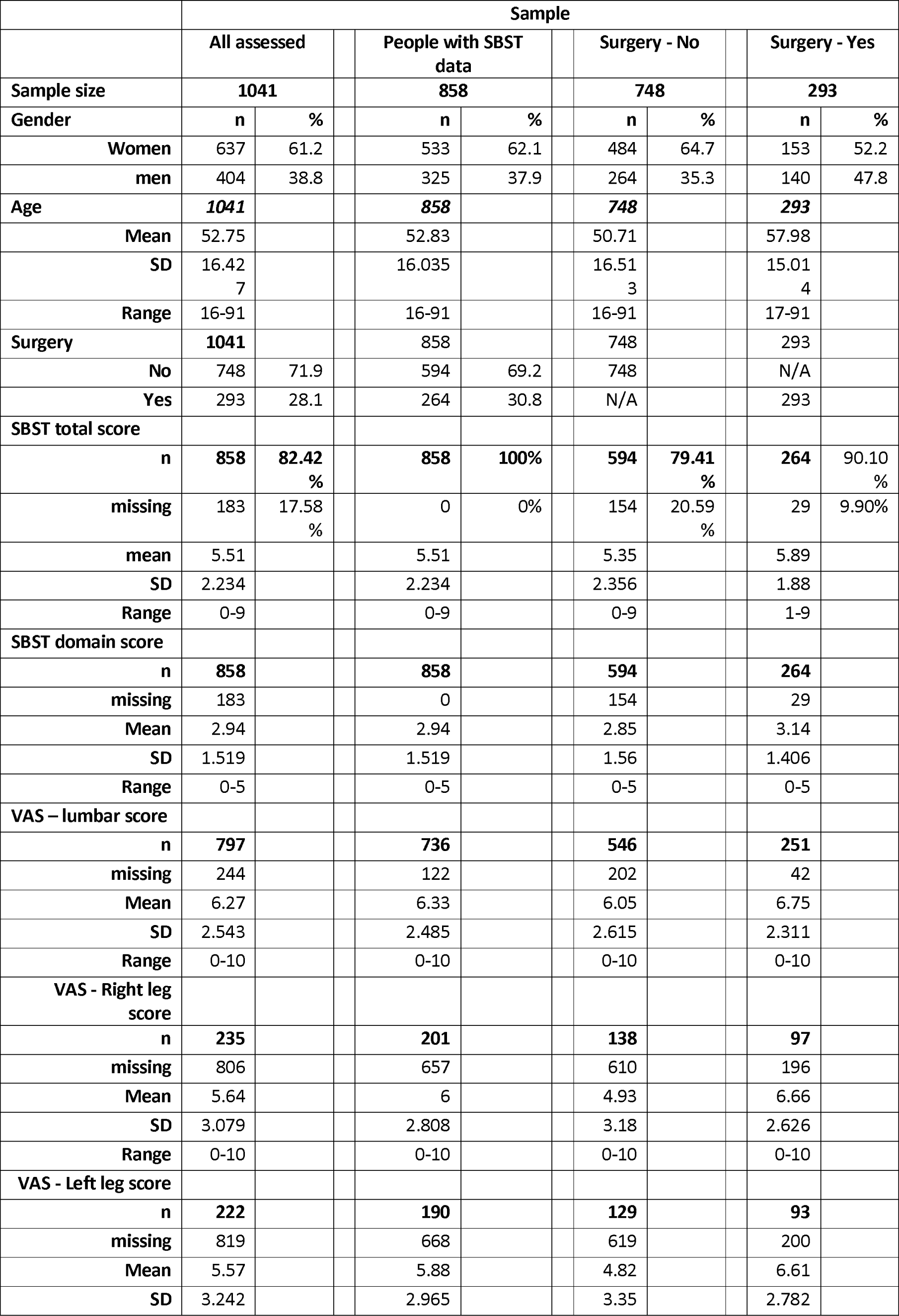
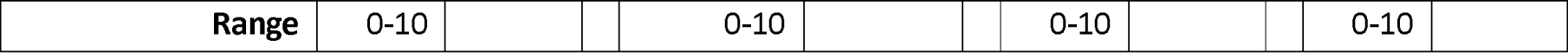
Characteristics, SBST and VAS summary scores total of four main samples

The subgroup with surgical lesions was older (58 v 51 yrs), had more males (48% v 35%), had greater pain scores for all 3 VAS’s [lumbar, right and left leg] done (6.8, 6.7, 6.6 v 6.1, 4.9, 4.8), and higher mean SBST total (5.9 v 5.4) and subscale (3.1 v 2.9) scores than the non-surgical group.

Tables 2 shows the distribution VAS scores of SBST total and SBST subscale scores and the distribution of (Table 3) for the four main samples. The profiles are relatively similar across these measures for the surgical and non-surgical samples. The surgical sample has lower percents with lower scores and higher percents with higher scores. The differences are relatively small.

**Table 3:** SBST total and domain score distributions of four main samples

| SBST Total Score | All |  |  | SBST scores |  |  | Surgery - No |  |  | Surgery - Yes |  |  |
| --- | --- | --- | --- | --- | --- | --- | --- | --- | --- | --- | --- | --- |
|  | n | % | Cum % | n | % | Cum % | n | % | Cum % | n | % | Cum % |
| **0** | 17 | 1.98 | 1.98 | 17 | 1.98 | 1.98 | 17 | 2.86 | 2.86 | 0 | 0.00 | 0.00 |
| **1** | 31 | 3.61 | 5.59 | 31 | 3.61 | 5.59 | 26 | 4.38 | 7.24 | 5 | 1.89 | 1.89 |
| **2** | 48 | 5.59 | 11.19 | 48 | 5.59 | 11.19 | 40 | 6.73 | 13.97 | 8 | 3.03 | 4.92 |
| **3** | 53 | 6.18 | 17.37 | 53 | 6.18 | 17.37 | 43 | 7.24 | 21.21 | 10 | 3.79 | 8.71 |
| **4** | 119 | 13.87 | 31.24 | 119 | 13.87 | 31.24 | 81 | 13.64 | 34.85 | 38 | 14.39 | 23.11 |
| **5** | 149 | 17.37 | 48.60 | 149 | 17.37 | 48.60 | 95 | 15.99 | 50.84 | 54 | 20.45 | 43.56 |
| **6** | 122 | 14.22 | 62.82 | 122 | 14.22 | 62.82 | 80 | 13.47 | 64.31 | 42 | 15.91 | 59.47 |
| **7** | 123 | 14.34 | 77.16 | 123 | 14.34 | 77.16 | 80 | 13.47 | 77.78 | 43 | 16.29 | 75.76 |
| **8** | 133 | 15.50 | 92.66 | 133 | 15.50 | 92.66 | 86 | 14.48 | 92.26 | 47 | 17.80 | 93.56 |
| **9** | 63 | 7.34 | 100.00 | 63 | 7.34 | 100.00 | 46 | 7.74 | 100.00 | 17 | 6.44 | 100.00 |
| **Total responses** | 858 | 82.42 % |  | 858 | 100 % |  | 594 | 79.41 % |  | 264 | 90.10 % |  |
| **Missing** | 183 | 17.58 % |  | 0 | 0 % |  | 154 | 20.59 % |  | 29 | 9.90 % |  |
| **Sample Total** | **1041** |  |  | **858** |  |  | **748** |  |  | **293** |  |  |
| **SBST Domain Score** | **n** | **%** |  | **n** | **%** |  | **n** | **%** |  | **n** | **%** |  |
| **0** | 72 | 8.39 | 8.39 | 72 | 8.39 | 8.39 | 61 | 10.27 | 10.27 | 11 | 4.17 | 4.17 |
| **1** | 82 | 9.56 | 17.95 | 82 | 9.56 | 17.95 | 58 | 9.76 | 20.03 | 24 | 9.09 | 13.26 |
| **2** | 173 | 20.16 | 38.11 | 173 | 20.16 | 38.11 | 124 | 20.88 | 40.91 | 49 | 18.56 | 31.82 |
| **3** | 194 | 22.61 | 60.72 | 194 | 22.61 | 60.72 | 124 | 20.88 | 61.78 | 70 | 26.52 | 58.33 |
| **4** | 172 | 20.05 | 80.77 | 172 | 20.05 | 80.77 | 119 | 20.03 | 81.82 | 53 | 20.08 | 78.41 |
| **5** | 165 | 19.23 | 100.00 | 165 | 19.23 | 100.00 | 108 | 18.18 | 100.00 | 57 | 21.59 | 100.00 |
| **Total responses** | 858 | 82.42 % |  | 858 | 100 % |  | 594 | 79.41 % |  | 264 | 90.10 % |  |
| **Missing** | 183 | 17.58 % |  | 0 | 0 % |  | 154 | 20.59 % |  | 29 | 9.90 % |  |
| **Sample Total** | **1041** |  |  |  |  |  | **748** |  |  | **293** |  |  |

## Discussion

Our results question using SBST to triage people with back pain in primary and secondary care. Over 80% of people with MRI identified surgical lesions considered to account for at least some of their symptoms were either at high or medium “risk”. Medium risk identifies individuals with unfavourable prognoses, high levels of physical prognostic indicators, appropriate for physiotherapy. High risk identifies individuals with very unfavourable prognoses, consistently high levels across psychosocial prognostic indicators, appropriate for management by a combination of physical and cognitive-behavioural approaches. All these people had surgical explanations for their symptoms.

The SBST simply indicates the extent of some impacts of back pain, higher scores indicate greater impact. It comes as no surprise to us that greater levels of pain have greater physical, mental and psychological health impacts than lesser levels of pain. However, a symptom severity questionnaire cannot, by definition, have diagnostic accuracy.

It is entirely appropriate for service providers to seek efficient ways of allocating limited resources. START back’s developers recognised a “one size fits all primary care strategy was suboptimum because it ignores the heterogenicity of patients”, and set out to, “test whether stratified care according to the estimated risk of poor prognosis (defined here as persistent disability because of back pain) improves clinical outcomes while remaining cost effective.”^5^

We have a different view. From our perspective the SBST represents a fundamental departure from standard accepted medical practice where diagnosis guides treatment. Our data show that neither the SBST risk groups, total scores nor subscale scores distinguish people who have structural lesions likely to be responsible for their pain, from those who do not. In our view, the established principle of making a diagnosis provides a “one size fits all” strategy which recognises patient heterogeneity and enables stratification.

Departing from the principle of basing treatment on diagnosis has significant implications raises important issues. In the absence of a diagnosis, it is not possible to collect robust evidence supporting the value of interventions prescribed for non-specific symptoms. There is a risk that ineffective management will be promoted and people for whom effective treatments are available will be missed or delayed. How correct is it to prescribe interventions without diagnoses? It is a reflection of the magnitude of the problem facing society that this largely untested approach features so prominently in national guidance (NICE and commissioning guidance produced by the BOA & SBNS)?

We consider that the accepted standard of care is to make a diagnosis on which to base treatment and prognosis. Challenges to this standard are encouraged but should be based on firm foundations. In their paper, claiming SBST’s efficacy, the developers provide a single reference supporting their statement that a “novel approach, gaining interest in other medical specialties, but not yet tested in the management of back pain, is to test whether stratified care according to the estimated risk of poor prognosis (defined here as persistent disability because of back pain) improves clinical outcomes while remaining cost effective.”^8^ This is one in a series of papers describing the use of prognostic models to “estimate the risk of future outcomes in individuals based on their clinical and non-clinical characteristics”. Whilst we agree that diagnosis is not be the only variable important in determining prognosis, it is hard to think it is not a principle factor.

We are not aware that such a prognostic model has been successfully applied to a symptom in the absence of a diagnosis. The SBST’s developers’ proposition is that prognosis can be estimated, and effective treatment prescribed from a symptoms severity questionnaire with no regard for diagnosis. It uses binary responses to non-specific questions including “I have had pain in the shoulder or neck at some time in the last 2 weeks” and “It’s not really safe for a person with a condition like mine to be physically active”. If it were possible to diagnose and prognosticate on the basis of such simple questions and answers it would certainly simplify healthcare provision. This might explaining its very early adoption by commissioning authorities. However, we are concerned that the rationale is flawed conceptually and supporting evidence is weak.

We believe a central problem is that it has become acceptable to label people suffering from lumbar spine related pain as suffering from “non-specific low back pain”. This term is then used as a diagnosis rather than a symptom. We consider that all pain will have an explanation and the best chance of helping people in pain is to understand the underlying cause, i.e, make a diagnosis. Our experience, and published work, shows that people with a surgically amenable structural explanation for pain are frequently not recognised because the symptoms of nerve root compression rarely conform to textbook and guideline descriptions or clinical expectations. In our published series of consecutive people undergoing surgery for lumbosacral disc disease, only 1 of 123 (0.81%) had a clinical presentation consistent with that documented in clinical guidances and textbooks.^9^ These factors are not consistent with diagnostic accuracy in primary care. We think similar scenarios probably apply to people with psychosocially or psychiatrically mediated symptoms for which more specific treatments may be suitable. For example, 80% of people with fibromyalgia referred for specialist psychiatric assessment were found to have a diagnosis.^10^

Lumbar pain is the final common pathway of multiple potential diagnoses. The vast majority will probably resolve spontaneously, and the cause is not important. However, for those with persistent disabling symptoms in whom we wish to prescribe treatments a diagnosis is required. Currently, we are unable to separate accurately the diagnoses of lumbar pain on clinical grounds, in part because this has not been studied carefully. The SBST measures severity across 9 ambiguous questions (e.g: “In general I have **not enjoyed** all the things I used to enjoy”). No one question is attributable to any one diagnosis. We have a choice: invest in understanding if it is possible to separate diagnosis accurately on clinical grounds in primary care; or, recognise that a diagnosis requires imaging and a specialist opinion so that treatable structural lesions are identified.

## Data Availability

The data is routinely collected clinical data which in principle we are happy to share. We have no consent to share but are investigating if sharing anonymised data is ethical.

